# The Impact of SARS-CoV-2 Variant to COVID-19 Epidemic in Brazil

**DOI:** 10.1101/2020.09.25.20201558

**Authors:** S. Celaschi

## Abstract

The impact of SARS-CoV-2 dominant global lineages to COVID-19 epidemics is for the first time modeled by an adaptation of the deterministic SEIR Model. Such a strategy may be applied worldwide to predict forecasts of the outbreak in any infected country. The objective of this study is to forecast the outcome of the epidemic in Brazil as a first cohort study case. The basic modeling design takes under consideration two of SARS-CoV-2 dominant strains, and a time-varying reproduction number to forecast the disease transmission behavior. The study is set as a country population-based analysis. Brazilian official published data from February 25 to August 30 2020 was employed to adjust a couple of epidemiological parameters in this cohort study. The population-based sample in this study, 4.2 Million Brazilians during the study period, is the number of confirmed cases on exposed individuals. Model parameters were estimated by minimizing the mean squared quadratic errors. The main outcomes of the study follows: The percentage values of non-symptomatic and symptomatic COVID-19 hosts were estimated to be respectively (54 ± 9) % and (46 ± 9) %. By the end of 2020, the number of confirmed cases in Brazil, within 95% CI, is predicted to reach 6 Million (5-7), and fatalities would account for 180.10^3^ (160–200).10^3^. Estimated forecast obtained preserving or releasing the NPIs during the last quarter of 2020, are included. Data points for extra three weeks were added after the model was complete, granting confidence on the outcomes. In 2020 the total number of exposures individuals is estimated to reach 13 ± 1 Million, 6.2% of the Brazilian population. Regarding the original SARS-CoV-2 form and its variant, the only model assumption is their distinct incubation rates. The variant SARS-CoV-2 form, as predicted by the SEIR adopted model, reaches a maximum of 96% of exposed individuals as previously reported for South America. By the end of 2020, a fraction in the range of 15–35 percentages of susceptible Brazilian individuals is to be depleted. Sufficient depletion of susceptibility (by NPIs or not) has to be achieved to weaken the global dynamics spread.

## Introduction

COVID-19 was first reported in Brazil in February 2020. One semester later, the country became one of the worst affected globally. Brazil comprises many states with vulnerable communities, and a relatively weak social protection system. After six months from the first reported case, the number of confirmed cases and deaths crossed 3.9 million and 120 thousand, respectively. Facts that question the availability of public health care for the sections of society that cannot afford private medical assistance.^**1,2**^ Since the start of the epidemic in Brazil, several types of Non-pharmaceutical Interventions - NPI have been adopted with varied success by the country’s 27 federal units and 5,596 municipalities. Virus transmission has dropped substantially in most units. However, by September 2020, the estimated reproduction number R remains above the unit.^3^ Thus, only mitigation (and not suppression) of the epidemic had been achieved so far. Closer surveillance of viral transmission quantifying the impact of different NPIs may help to minimize future infections. Moreover, continued monitoring of the genetic diversity of the virus lineages circulating globally became necessary. As recently suggested, virus diversity is of prime importance in the SARS-CoV-2 transmission.^4,5^ By the end of February 2020, before the implementation of NPIs and travel bans, different SARS-CoV-2 lineages had emerged in Brazil from Europe.^6^ In the beginning, the epidemic had spread mostly locally and within-state borders.^6^

The objective of this study is to model the outcome of the epidemic in Brazil as a first cohort study case. The basic modeling design takes under consideration two of SARS-CoV-2 dominant strains, and a time-varying reproduction number to forecast the disease transmission behavior. The study is set as a country population-based analysis employing Brazilian official published data for the period February 25 to August 30. 2020. The population-based sample in this study, 4,238,446 Brazilians, is the number of confirmed cases on exposed individuals. Model parameters were estimated by minimizing the mean squared quadratic errors. Epidemiological models are commonly stochastic, network-based, spatially diffusive, using meta-population dynamics.^7,8,9^ However, in the interpretation of physical processes, the parameters of deterministic models are directly related.^10^ As a drawback, deterministic models impose restrictive analysis, once the dynamics of the host population has free will and the virus undergoes favorable mutations upon positive selection.^5^ Host dynamics was account for into the model by spreading the use of time-varying reproduction number.^11^ Regarding the pathogen, the variant SARS-CoV-2 D614G mutation, which became globally proposed as dominant, was incorporated to the model.^5^ In short, the methodology employed is an adaptation to the SEIR Model, which takes into account two lineages of the SARS-CoV-2, and a time-varying reproduction number to account for changes in the dynamics of the COVID-19 transmission behavior. Such a methodology can be applied worldwide to predict forecasts of the outbreak in any infected country.

### Increased SARS-CoV-2 Variant Frequency on Global Distribution, and Prevalence of Non-symptomatic Infections

The persistence of the SARS-CoV-2 pandemic built an accumulation of relevant mutations, even though its diversity is reported to be low.^12^ Increasing frequency of spike amino acid variant has been reported globally^5,13^ The spike protein determines the infectivity of the virus, and its transmissibility in the host. Mutations in the gene encoding Spike (S) protein are being reported since SARS-CoV-2 was identified in humans. Those mutations of S proteins affect viral life cycle and its interaction with the host. Very recent studies suggest that the observed increased transmission reported in Europe and Americas may be associated with the most dominant variant.^5^ Experimental evidence has been that the so called D614G spike variant is associated with greater infectivity as well as higher viral loads.^5^ Global tracking data show that the G614 variant in Spike has spread faster than D614 comparing two time periods apart by a 2-week gap. The fraction for South America prior to March 1 are D = 38%, G = 62%, and D = 4%, G = 96% in the period March 2020, 21-30. They also show running weekly average counts of sampled sequences exhibiting the D614 and G614 variants for South America between January 12 and May 12. The results are explained assuming the virus being more infectious. On the other hand, the authors did not find evidence of G614 effects on disease severity. As recently reported elsewhere, the Spike protein 614 polymorphism has little variance in growth rates among clusters and presented no significant difference in initial growth rates.^13^ Those facts are incorporated in our model and evidences are demonstrated.

SARS-CoV-2 has shown to be surprisingly harmless to a largest fraction of infected individuals, heavily harm to many others, and lethal to a few percent of exposed hosts. Infected persons who remain asymptomatic and pre-symptomatic (here on named non-symptomatic) play a significant role in the ongoing COVID-19 pandemic. Therefore, the prevalence of non-symptomatic SARS-CoV-2 infected host has to be incorporated in modeling any dynamics of COVID-19 outbreak. Applying statistical analysis on recent published and reliable data, the percentage value of non-symptomatic and symptomatic hosts were estimated to be respectively (54 ± 9) % and (46 ± 9) %.^14^

### Serial Intervals and Inference of time-varying reproduction number

Serial interval (SI) is an essential metric for estimating main epidemiological parameters, which in turn are used to predict disease trends, interventions and health care demands. SI depends on the pathogen incubation period which quantifies the biological process of relevant virus mutation, disease progression and tends to follow distributions resulting from genetic differences.^15^ Variations in SI can occur and may have significant implications for the transmission dynamics affecting other epidemic parameters. The real-time transmissibility of an infectious disease is often characterized by the instantaneous reproduction number *R(t)* defined as the expected number of secondary infections caused by an infector within a short time window. Equivalently, *R(t)* can be expressed as the transmission rate *β(t)* divided by the rate γ_*o*_ into which infected people recover or die. Mitigation policies aim to control the outbreak reducing the *R(t)* value. A number of methods are available to estimate effective reproduction numbers during epidemics.^16^ With this aim, a method for estimating *R(t)* using branching processes was developed.^17^ It relies on two inputs: a disease incidence time series (the numbers of new observed cases at successive times) and an estimate of the distribution of SI. More recently, this statistical framework was extended to allow adding data on known pairs of index and secondary cases from which the serial interval is directly estimated.^11^ During the COVID-19 outbreak in Brazil, *R(t)* was estimated following the procedure available on-line.^18^

### Estimative of Infected Individuals and Fatal Outcomes

There are two measures used to assess the proportion of infected individuals with fatal outcomes, the infection fatality ratio (IFR), which estimates this proportion of deaths among all infected individuals, and case fatality ratio (CFR), which estimates this proportion of deaths among identified confirmed cases. Under estimations of the rate of disease transmission spoils the calculus of IFR.^19^ A significant proportion of infected people are undetected because they are non-symptomatic, and fail to present at healthcare facilities. Furthermore, testing capacity has been limited in Brazil, and restricted to people with severe cases and priority risk groups. To accurately measure IFR, the complete number of infections, and deaths caused by the disease must be known. Very recently, a careful evaluation of the IFR was reported to the COVID-19 outbreak in Brazil.^20^ A country-wide average IFR of 1.05% (95% CRI: 0.96–1.17%) was reported by the authors. On the other hand, reliable CFRs are generally obtained at the end of an outbreak, after most infected individuals either died or recovered. CFR calculated using the above relation during ongoing epidemics is influenced by lags in report dates for cases and deaths. Obtaining both IFR and CFR from available data allows estimating the fraction of symptomatic hosts in order to apply to the model.

### Modeling COVID-19 Pandemics in Brazil

Epidemiological models are useful in estimating main parameters of a pathogen spread like SARS-CoV-2. Such models may evaluate the efficacy of specific interventions, identify relevant strategies, and forecast future scenarios.^21^ Data fitting to adjust model parameters provide estimative on the basic and effective reproduction numbers, the case fatality ratio (CFR), the infection fatality ratio (IFR), among others. Parameterized models may clarify ambiguities create by non-symptomatic infected hosts and delays between incubation, infection and heal/death. Important to note that official reported confirmed COVID-19 cases usually miss non symptomatic infections, which can bias estimates of disease severity and IFR, so epidemiological models could help reduce this uncertainty. The Susceptible-Exposed-Infected-Removed - SEIR model was adapted in this study to take into account the original SARS-Cov-2 D-form and its dominant G-variant with their own incubation rates, a pre-estimated fraction of symptomatic hosts, and a pre-inferred time-varying reproduction number. COVID-19 has a latent or incubation period, during which individuals are said to be infected but not infectious. Members of this population in latent stage are labeled as Exposed (but not infectious). Taken into consideration the original SARS-Cov-2 D-form and its dominant G-variant labeled as D and G, the deterministic model with the groups: Susceptible, Exposed (D and G), Infected (D and G), and Removed (recovered and deaths/fatalities) is labeled as the so called SE_D_E_G_I_D_I_G_R Model. The number of fatalities is assumed dependent on the confirmed COVID-19 cases (Infected D and G). This dependence is not linear neither monotonic, and is obtained from official reported cumulative values. COVID-19 cases in Brazil are reported by public health and private services, and interrelated on a website which summarizes daily the aggregated counts.^22^ Worth to mention that by the time this study was conducted, reliable estimative on the number of sub notifications and information on the number of non-symptomatic were unreliable. Under those assumptions, the set of Ordinary Differential Equations – ODE governing our SE_D_E_G_I_D_I_G_R model follows;

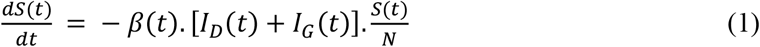

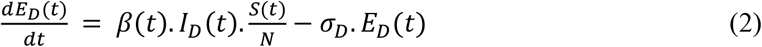

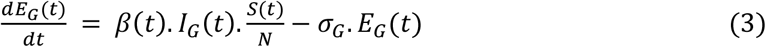

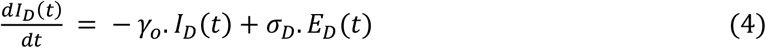

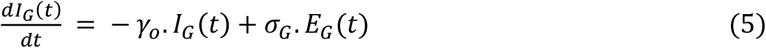

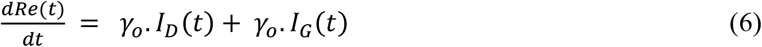

where *S(t), E*_*D*_*(t), E*_*G*_*(t), I*_*D*_*(t), I*_*G*_*(t), and Re(t)*, are respectively daily numbers of Susceptible, Exposed (D and G), Infected (D and G), Removed (recovered and deaths) individuals. *S(t) + E*_*D*_*(t) + E*_*G*_*(t) + I*_*D*_*(t) + I*_*G*_*(t) + Re(t) = N =* Constant. *β(t)= R(t)/*γ_*o*,_ *where R(t)* is the time-varying reproduction number. *R(t=0)=R*_*o*_.γ_*o*_ is the removed rate for which infected individuals (symptomatic and non symptomatic) recover or die, leaving the infected groups *I*_*D*_*(t) and I*_*G*_*(t)*. The accumulated SARS-Cov-2 confirmed cases are obtained from:

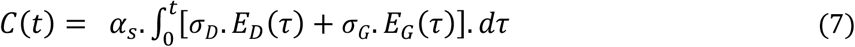

where α_*S*_ = 0.47 ± 0.09 is the estimated fraction of symptomatic individuals. Major assumptions for this model are respectively the incubation rates: *σ*_*D*_ *= 0*.*5* and *σ*_*G*_ *= σ*_*D*_*/2* for D and G SARS-CoV-2 exposed hosts. It is assumed that both D and G groups share the same *R(t)* function. In short, *N,σ*_*D*_, and *σ*_*G*_ are the only fitting parameters to data. The susceptible number *N=60* Million (28.7% of the Brazilian population) is chosen as the minimum number of susceptible hosts to account for the accumulated confirmed cases in the study period. Fatalities were modeled as a function of the confirmed cases *C(t)*, and this dependence, obtained from official reported cases and deaths, is not linear neither monotonic as discussed latter.

## Results and Discussion

Official Brazilian data from February 29 to August 19 2020 (EPI week # 9), was considered in order to estimate the main parameters that govern the dynamics established by Eqns. (1) to (7).^22^ All model parameters were estimated by minimizing the mean squared quadratic errors. A key parameter in deterministic transmission models is the basic reproductive number *R*_*o*_, which is quantified by both, the pathogen and the susceptible population in which it circulates. The methodology to estimate *R*_*o*_ was presented in a previous publication.^16^ When infection is spreading through a population that may be partially non-symptomatic, besides *R*_*o*_, an accurate estimation of the *R(t)* value is crucial to plan and control the infection.^11^ In this work, *R(t)* was estimated from a Gamma Distribution Time Generations (Gamma Mean Value = 3.6, Variance = 4.8) on confirmed cases from February 25 to August 19.^16^ Table 1 presents some estimated values obtained from a 7 days running window.

**Table 1.** Selected time-varying reproduction values estimated from a Gamma Distribution Time Generation (Mean Value = 3.6, Variance = 4.8) applied on a 7 days running window of confirmed cases.^18^

| Start time | Avr. time | End time | Mean (R) | Std (R) | Q 0.025 (R) | Q 0.05 (R) | Q 0.25 (R) | Median (R) | Q 0.75 (R) | Q 0.95 (R) | Q 0.975 (R) |
| --- | --- | --- | --- | --- | --- | --- | --- | --- | --- | --- | --- |
| 2 | 5 | 8 | 2.24 | 0.19 | 1.89 | 1.94 | 2.11 | 2.24 | 2.37 | 2.56 | 2.63 |
| 9 | 12 | 15 | 2.28 | 0.14 | 1.9 | 1.94 | 2.07 | 2.17 | 2.26 | 2.41 | 2.46 |
| 16 | 19 | 22 | 1.97 | 0.06 | 1.87 | 1.89 | 1.95 | 2 | 2.04 | 2.1 | 2.12 |
| 23 | 26 | 29 | 1.55 | 0.02 | 1.87 | 1.88 | 1.9 | 1.92 | 1.93 | 1.95 | 1.96 |
| 30 | 33 | 36 | 1.45 | 0.01 | 1.45 | 1.45 | 1.46 | 1.47 | 1.47 | 1.48 | 1.49 |
| 37 | 40 | 43 | 1.34 | 0.01 | 1.42 | 1.42 | 1.43 | 1.43 | 1.43 | 1.44 | 1.44 |
| 44 | 47 | 50 | 1.27 | 0 | 1.32 | 1.32 | 1.32 | 1.33 | 1.33 | 1.33 | 1.33 |
| 51 | 54 | 57 | 1.23 | 0 | 1.26 | 1.26 | 1.27 | 1.27 | 1.27 | 1.27 | 1.27 |
| 58 | 61 | 64 | 1.24 | 0 | 1.23 | 1.23 | 1.23 | 1.24 | 1.24 | 1.24 | 1.24 |
| 65 | 68 | 71 | 1.23 | 0 | 1.23 | 1.23 | 1.23 | 1.23 | 1.24 | 1.24 | 1.24 |
| 72 | 75 | 78 | 1.2 | 0 | 1.22 | 1.22 | 1.22 | 1.22 | 1.22 | 1.22 | 1.22 |
| 79 | 82 | 85 | 1.2 | 0 | 1.2 | 1.2 | 1.2 | 1.2 | 1.2 | 1.2 | 1.2 |
| 86 | 89 | 92 | 1.17 | 0 | 1.19 | 1.19 | 1.19 | 1.19 | 1.19 | 1.19 | 1.19 |
| 93 | 96 | 99 | 1.16 | 0 | 1.17 | 1.17 | 1.17 | 1.17 | 1.17 | 1.17 | 1.17 |
| 100 | 103 | 106 | 1.13 | 0 | 1.15 | 1.15 | 1.15 | 1.15 | 1.15 | 1.15 | 1.15 |
| 107 | 110 | 113 | 1.11 | 0 | 1.12 | 1.12 | 1.12 | 1.12 | 1.12 | 1.12 | 1.12 |
| 114 | 117 | 120 | 1.11 | 0 | 1.11 | 1.11 | 1.11 | 1.11 | 1.11 | 1.11 | 1.11 |
| 121 | 124 | 127 | 1.09 | 0 | 1.11 | 1.11 | 1.11 | 1.11 | 1.11 | 1.11 | 1.11 |
| 128 | 131 | 134 | 1.08 | 0 | 1.09 | 1.09 | 1.09 | 1.09 | 1.09 | 1.09 | 1.09 |
| 135 | 138 | 141 | 1.07 | 0 | 1.08 | 1.08 | 1.08 | 1.08 | 1.08 | 1.08 | 1.08 |
| 142 | 145 | 148 | 1.07 | 0 | 1.07 | 1.07 | 1.07 | 1.07 | 1.07 | 1.07 | 1.07 |
| 149 | 152 | 155 | 1.06 | 0 | 1.07 | 1.07 | 1.07 | 1.07 | 1.07 | 1.07 | 1.07 |
| 156 | 159 | 162 | 1.06 | 0 | 1.06 | 1.06 | 1.06 | 1.06 | 1.06 | 1.06 | 1.06 |
| 163 | 166 | 169 | 1.05 | 0 | 1.06 | 1.06 | 1.06 | 1.06 | 1.06 | 1.06 | 1.06 |
| 170 | 173 | 176 | 1.04 | 0 | 1.05 | 1.05 | 1.05 | 1.05 | 1.05 | 1.05 | 1.05 |
| 177 | 180 | 183 | 1.04 | 0 | 1.04 | 1.04 | 1.04 | 1.04 | 1.04 | 1.04 | 1.04 |

Fig. 1 presents (right-vertical axis) the mean time-varying R(t) (data from Table 1 - solid blue diamond)^11^, vertical bars represent 5 to 95% CRI. Time *t* is given in units of epidemiological weeks, commonly referred to as EPI weeks. The first two weeks of epidemic spread in Brazil presented, *R(t)* values larger than 2. However, after two weeks, *R(t)* shows a monotonic decline, reaching, after 17 weeks, values slightly above the unit, as a consequence of NPI federal/regional interventions, and collective acquired immunity across country (Fig.1). NPIs in Brazil were implemented between EPI week #10 and #12 across the countries’ 27 federal states, and consisted of social distancing, mask wearing, work at home, school, and stores closures.^6, 16^ Figure 1 also presents for *R(t > 9 + 27 EPI)* estimated forecasts (shaded black area) preserving or releasing the NPIs during the last quarter of 2020.

**Figure 1.**
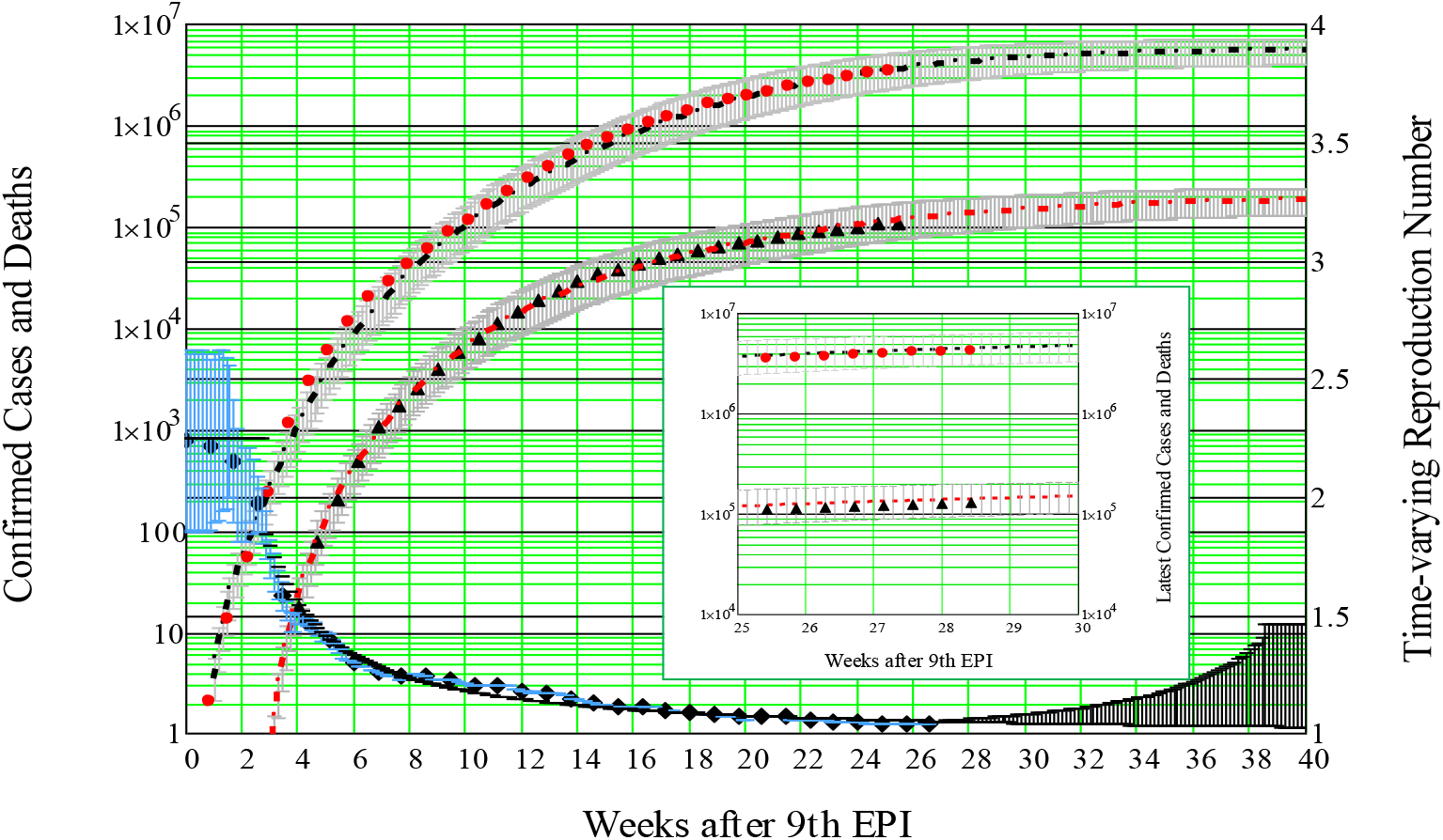
Data on confirmed cases and fatalities are shown by solid red circles, and solid black triangles respectively. Error bars account for the RMS values on the SEIR parameters. The fitting values to SEIR model are: N = 6.10^7^, β_o_ = 1.265, σ_D_ = 0.25, σ_G_ = 2σ_D_. R_o_=2.25 and α_s_= 0.47 ± 0.09. Average values and error bars (vertical in black) for infected individuals, and fatalities are shown respectively in solid red circles and solid black triangles. The mean fitting to the infected individuals (logarithmic scale) presents standard deviation SD = 0.08. Right vertical axis shows: the mean time-varying R(t) (data from Table 1 - solid blue diamond)^11^, vertical bars represent 5 to 95% CRI; (b) R(t > 9 + 27 EPI) estimated forecasts (shaded black area) preserving or releasing the NPIs. Data points for the last three weeks (Figure Inset - solid circles and triangles) were added after the model was complete, granting confidence on the outcomes.

Assuming to the end of 2020 the preservation of all NPIs, as social distancing, mask wearing, gloves, hand wash, among others, a first fitting function is selected to model a continuous approach of *R(t)* to its unit value. This first fitting is shown in Fig.3 as a dash-and-dot black trace. On a different scenario, aiming to model the second COVID-19 wave spreading in Brazil during the second half of 2020, a second fitting is proposed. This second fitting is included in Fig. 3 as a dash red trace. In this scenario, a progressive release of all NPI is adopted leading *R(t)* to return to 65% of its initial value R_o_. Both scenarios will be discussed latter.

Furthermore, fittings to data on accumulated confirmed cases (symptomatic infected individuals), according to Eqn. 7 are presented in Fig. 1. The only fitting values are: *N =6×10*^*7*^ (28.7% of Brazilian population), β*(t=0) = 1*.*265*, σ_*D*_ *= 0*.*25*, σ_*G*_ *= 2*σ_*D*_. Initial conditions: S(0) + E(0) = N, E(0) = E_D_(0) + E_G_(0) = 8, I_D_(0) = I_G_(0) = R_e_(0) = 0. The SARS-Cov-2 spike variant G associated with higher viral loads is modeled by a higher (two fold) incubation rate ∫_*G*_ as compared to ∫_*D*_. The fraction of symptomatic hosts within 95% CRI was estimated comparing IFR to CFR reported values.^20^ The estimated mean value and deviation are consistent with previous analysis.^14^ Fatalities were modeled as a function of the confirmed cases *C(t)*, and the result is included in Figure 1. By the end of 2020, the predicted numbers of confirmed cases in Brazil, within 95% credible intervals, may reach 6 Million (5-7) and fatalities would account for 180 thousand (160–200). The total number of infected individuals is estimated to reach (13 ± 1) Million, 6.2% of the population.

Fatalities as a function of the confirmed cases *C(t)*, present a time-dependence that is not linear neither monotonic. As shown in Fig. 2, the non-zero ratio of deaths/(confirmed cases) by COVID-19 (open red circles), begins three weeks after the first reported case, rises monotonically, after roughly seven weeks reaches a maximum of 7%, and drops to a ratio around 2.6%. Possible explanations are: NPIs as social distancing; the increased number of tests and reports available after the first weeks of the epidemic outbreak; and ICU improved medical procedures. COVID-19 was detected in Brazil in the 9th EPI week of 2020, and testing procedures for the SARS-CoV-2 virus was effectively included in the surveillance four weeks later. Furthermore, ventilators were scarce at the first weeks. The fitting (dashed blue curve) to the experimental data multiplied by *C(t)* (Eq.7) allows estimating the accumulated fatalities as presented in Fig. 1.

**Figure 2.**
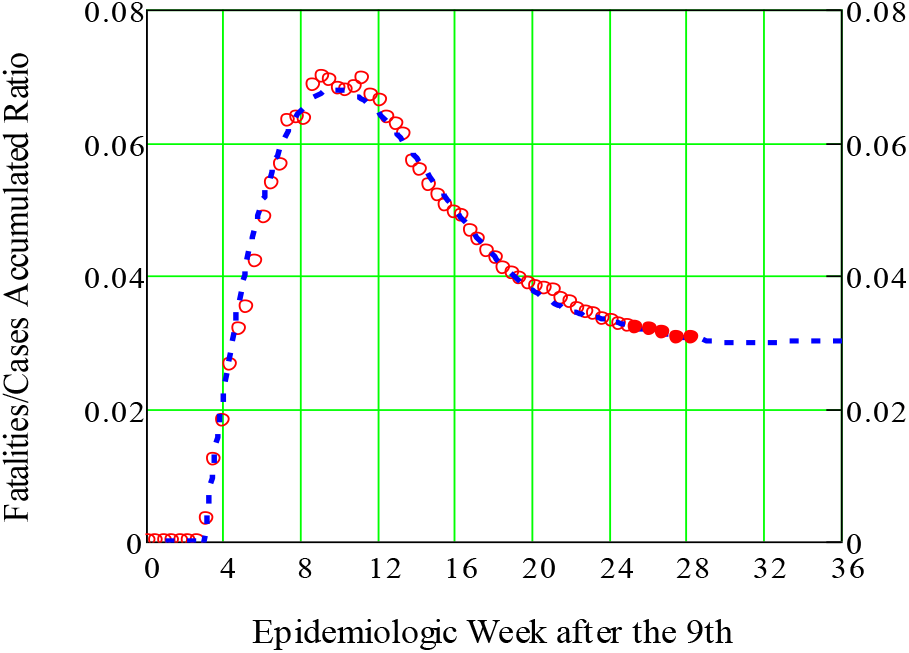
Time dependence of the ratio Fatalities/(Confirmed Cases). This ratio presents a time-dependence that is not linear neither monotonic. The non-zero ratio of deaths/(confirmed cases) by COVID-19 (open red circles), begins three weeks after the first reported case, rises monotonically, after roughly seven weeks reaches a maximum of 7%, and drops to a ratio around 2.6 %. The fitting equation (dashed blue curve) to the experimental data multiplied by Eq.7 allows estimating the accumulated fatalities as shown in Fig. 1. Data points for the last three weeks (solid red circles) were added after the model was complete, granting confidence on the outcome.

Regarding the original SARS-CoV-2 D-form and its G-variant, the only distinction in modeling is their own incubation rates. Additionally, it is assumed that both forms produce the same disease fatalities, and share the same instantaneous reproductive number.^13^ The initial percentage values set for individuals exposed to the D-form and its G-variant were respectively 62% and 38%.^4^ Figure 3 presents the evolution of exposed and infected host to the original D-form and its G-variant according to model. These results confirm the prevalence of the G-variant form in COVID-19 pandemic as globally predicted. The G-variant form reaches a maximum of 96% of exposed individuals as previously reported for South America.^4^ The resulted shift in the epidemiological spread suggests that the G-variant form may have a fitness advantage. These findings support continuing surveillance of SARS-CoV-2 mutations to support development of immunological interventions.

**Figure 3.**
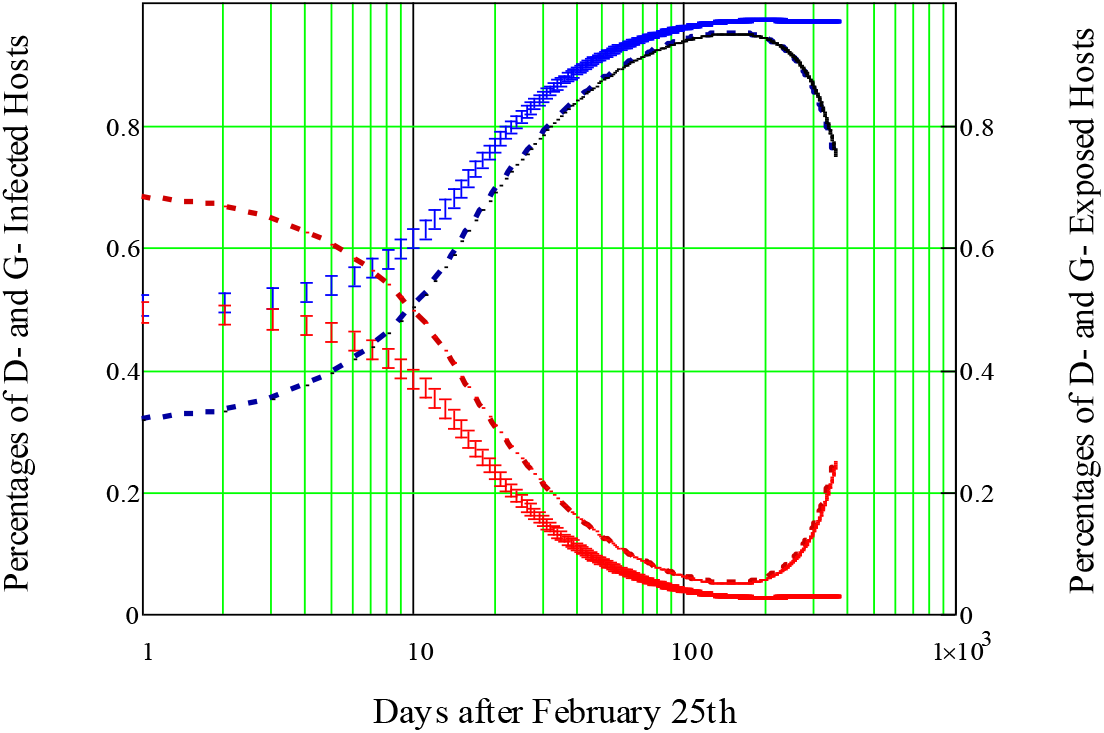
Estimated evolution of exposed and infected host to SARS-CoV-2 original D-form and G-variant. Broken red and blue lines present respectively the evolution of D and G exposed hosts to SARS-CoV-2. Vertical red and blue bars are respectively the evolution α_S_ = 0.47 ± 0.09 and E(0)= 6 ± 2) of D and G infected hosts. According to model, the percentage of exposed hosts to prevalent G-form of SARS-CoV-2 quickly rises and reaches a maximum of 96% as previously reported for South America.

Latin America became an epicenter of the COVID-19 pandemic in May 2020, driven by Brazil’s exponentially risen confirmed cases as the number of known infections in Europe fell. COVID-19 was first reported in Brazil in February 2020. In early June 2020, Brazil began averaging about 1,000 deaths per day from Covid-19, joining the United States as the countries with the world’s largest death tolls. Then, starting in August, the spread of the virus reduced, and the daily death toll began to drop. In a short period of time, shopping malls, restaurants and beaches started to draw crowds again. Tourist attractions reopened in several cities, and case numbers started rising and daily death tolls shot above 500 a day again. Major cities such as São Paulo, Rio de Janeiro, and Recife found hospital bed occupation rates reaching over 80 percent, and pressure increased on Brazilian authorities to re-establish restrictive measures. In short, during second half of 2020 a second wave is spreading fast in Brazil as in Europe, and across other continents.

Figure 7 presents daily evolution of symptomatic infected host to the original D-form and its G-variant according to model estimative. These results confirm the prevalence of the G-variant form in COVID-19 pandemic as globally predicted, and reported for South America. They also point out to the presence of a second outbreak by the end of 2020 as a result of the release of NPIs across the country. These previsions suggest reinforcing the NPIs, and continuing the surveillance of SARS-CoV-2 mutations to support development of immunological interventions.

**Figure 7.**
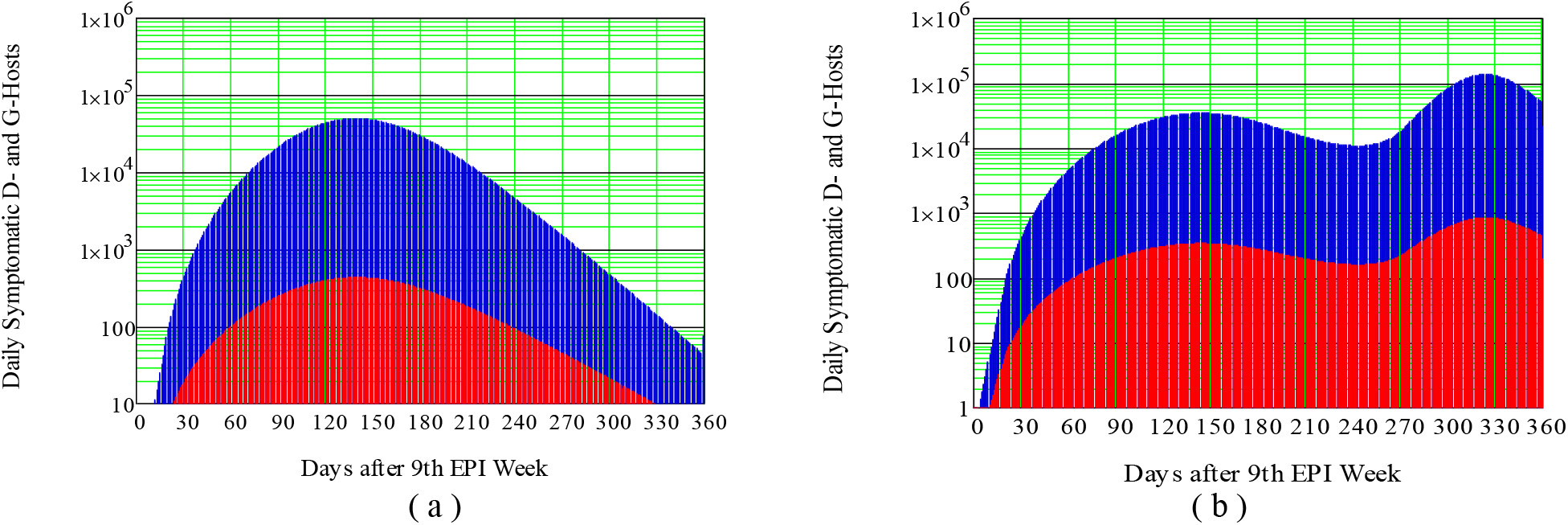
Estimated evolutions of symptomatic infected host to SARS-CoV-2 original D-form and G-variant are presented in (a) and (b). Red and blue shades present respectively the evolution of D and G exposed hosts to SARS-CoV-2. (a) Preserving NPIs. (b) Progressive release of NPIs as reported world wise during the last quarter of 2020, and as modeled in Fig.1.

## Conclusions

COVID-19 dynamics was modeled by an adaptation of the SEIR model, taking into consideration two lineages of the SARS-CoV-2, and a time-varying reproduction number. Such a methodology can be applied worldwide to predict forecasts of the outbreak in any infected country. The epidemic in Brazil was selected as a first study case. The estimative on the accumulated number of infected individuals, and fatalities agrees with reported values (5–95% CRI) for orders of magnitude. By the end of 2020, the predicted numbers of confirmed cases in Brazil will reach (6 ± 1) Million (5–95% CRI) and fatalities would reach (180 ± 20) thousand. The total number of infected individuals is estimated to reach (13 ± 1) Million, 6.2% of the population by the end of 2020. The variant SARS-CoV-2 form reaches a maximum of 96% of exposed individuals. An important conclusion worth to point out follows. Despite the social and mental commotion that COVID-19 has imposed so far, most of the Brazilian and the world populations are still susceptible to SARS-CoV-2 infection. By the end of 2020, a fraction in the range of 15–35 percentages of susceptible Brazilian individuals is predicted to be depleted. Sufficient depletion of susceptibility (by NPIs or not) has to be achieved to weaken the global dynamics spread.

## Data Availability

All data referred to in the manuscript is already published and the sources are added to manuscript reference section. All relevant ethical guidelines have been followed.

https://covid.saude.gov.br/

## Acknowledgments

The author acknowledges the financial support from FUNTTEL. Financial grant # 01.16.0053.01 FINEP/MCTI received from the Brazilian Ministry of Science, Communications, and Innovation. This paper reflects only the author’s views and the Agencies are not responsible for any use that may be made of the information contained therein. Thanks to Dr Kiyoshi Yoneda and Dr Roberto Panepucci for enlighten suggestions.

## Notes

### Competing Interest Statement

The authors have declared no competing interest.

### Funding Statement

This work was granted by the financial support from FUNTTEL. Financial grant # 01.16.0053.01 FINEP/MCTI from the Brazilian Ministry of Science Communications and Innovation.

### Summary of Updates

The impact of SARS-CoV-2 dominant global lineages to COVID-19 epidemics is for the first time modeled by an adaptation of the deterministic SEIR Model. Such a strategy may be applied worldwide to predict forecasts of the outbreak in any infected country. The objective of this study is to forecast the outcome of the epidemic in Brazil as a first cohort study case. The basic modeling design takes under consideration two of SARS-CoV-2 dominant strains, and a time-varying reproduction number to forecast the disease transmission behavior. The study is set as a country population-based analysis. Brazilian official published data from February 25 to August 30 2020 was employed to adjust a couple of epidemiological parameters in this cohort study. The population-based sample in this study, 4.2 Million Brazilians during the study period, is the number of confirmed cases on exposed individuals. Model parameters were estimated by minimizing the mean squared quadratic errors. The main outcomes of the study follows: The percentage values of non-symptomatic and symptomatic COVID-19 hosts were estimated to be respectively (54 +/− 9) % and (46 +/− 9) %. By the end of 2020, the number of confirmed cases in Brazil, within 95% CI, is predicted to reach 6 Million (5-7), and fatalities would account for 180.103 (160-200).103. Estimated forecast obtained preserving or releasing the NPIs during the last quarter of 2020, are included. Data points for extra three weeks were added after the model was complete, granting confidence on the outcomes. In 2020 the total number of exposures individuals is estimated to reach 13 +/− 1 Million, 6.2% of the Brazilian population. Regarding the original SARS-CoV-2 form and its variant, the only model assumption is their distinct incubation rates. The variant SARS-CoV-2 form, as predicted by the SEIR adopted model, reaches a maximum of 96% of exposed individuals as previously reported for South America. By the end of 2020, a fraction in the range of 15-35 percentages of susceptible Brazilian individuals is to be depleted. Sufficient depletion of susceptibility (by NPIs or not) has to be achieved to weaken the global dynamics spread.

