## Supplementary Material for "The Impact of SARS-CoV-2 Variant to COVID-19 Epidemic in Brazil"

*S. Celaschi*
*CTI Renato Archer, Rod. D Pedro I (SP-65), Km 143,6 - CEP 13069-901, Campinas – SP, 13069, Brazil.*

*September, 2020*

Latin America became an epicenter of the COVID-19 pandemic in May 2020, driven by Brazil's exponentially risen confirmed cases as the number of known infections in Europe fell. COVID-19 was first reported in Brazil in February 2020. In early June 2020, Brazil began averaging about 1,000 deaths per day from Covid-19, joining the United States as the countries with the world's largest death tolls. Then, starting in August, the spread of the virus reduced, and the daily death toll began to drop. In a short period of time, shopping malls, restaurants and beaches started to draw crowds again. Tourist attractions reopened in several cities, and case numbers started rising and daily death tolls shot above 500 a day again. Major cities such as São Paulo, Rio de Janeiro, and Recife found hospital bed occupation rates reaching over 80 percent, and pressure increased on Brazilian authorities to re-establish restrictive measures. In short, during last quarter of 2020 a second wave is spreading fast in Brazil as in Europe, and across other continents. Since the start of the epidemic in Brazil, several types of Non-pharmaceutical Interventions - NPI have been adopted with varied success by the country's 27 federal units and 5,596 municipalities. However, by August 2020, the estimated time-varying reproduction number *R(t)* still remains above the unit[1]**.** Thus, only mitigation (and not suppression) of the epidemic had been achieved so far.

By the end of February 2020, before the implementation of domestic NPIs, different SARS-CoV-2 lineages had emerged in Brazil from Europe. The objective of this study is to model the outcome of the epidemic in Brazil as a first cohort study case. The methodology employed - an adaptation to the well known SEIR Model – is described in details in this supplementary material in order to account for changes in the dynamics of the COVID-19 transmission behavior. Such a methodology can be applied worldwide to predict forecasts of the outbreak in any infected country.

#### Modeling COVID-19 in Brazil

Epidemiological models are commonly stochastic, network-based, spatially diffusive, using meta-population dynamics.[2,3,4] However, in the interpretation of physical processes, the parameters of deterministic models are directly related.[10>5] The Susceptible-Exposed-Infected-Removed - SEIR model was adapted in this study. The model takes into account the original SARS-CoV-2 D-form, and its dominant G-variant[6.7.8] with their respective incubation rates, a pre-estimated fraction of symptomatic hosts, and a pre-inferred time-varying reproduction number[9]. COVID-19 has a latent or incubation period, during which individuals are said to be infected but not infectious. Members of this population in latent stage are labeled as Exposed (but not infectious). Taken into consideration the original SARS-Cov-2 D-form and its dominant G-variant labeled as D and G, the deterministic model with the groups: Susceptible, Exposed (D and G), Infected (D and G), and Removed (recovered and deaths/fatalities) is labeled as $SE_DE_GI_DI_GR$ Model. The number of fatalities is assumed dependent on the confirmed COVID-19 cases (Infected D and G). This dependence is not linear neither monotonic, and is obtained from official reported cumulative values. Under those assumptions, the set of Ordinary Differential Equations – ODE governing our $SE_DE_GI_DI_GR$ model follows;

$$\frac{dS(t)}{dt} = -\beta(t).[I_D(t) + I_G(t)].\frac{S(t)}{N} \quad (1)$$

$$\frac{dE_D(t)}{dt} = \beta(t).I_D(t).\frac{S(t)}{N} - \sigma_D.E_D(t) \quad (2)$$

$$\frac{dE_G(t)}{dt} = \beta(t).I_G(t).\frac{S(t)}{N} - \sigma_G.E_G(t) \quad (3)$$

$$\frac{dI_D(t)}{dt} = -\gamma_o.I_D(t) + \sigma_D.E_D(t) \quad (4)$$

$$\frac{dI_G(t)}{dt} = -\gamma_o.I_G(t) + \sigma_G.E_G(t) \quad (5)$$

$$\frac{dRe(t)}{dt} = \gamma_o.I_D(t) + \gamma_o.I_G(t) \quad (6)$$

where $S(t), E_D(t), E_G(t), I_D(t), I_G(t),$ *and* $Re(t),$ are respectively daily numbers of Susceptible, Exposed (D and G), Infected (D and G), Removed (recovered and deaths) individuals. $S(t) + E_D(t) + E_G(t) + I_D(t) + I_G(t) + Re(t) = N$ = Constant. $\beta(t)= R(t)/\gamma_o$, *where* $R(t)$ is the time-varying reproduction number. $R(t=0)=R_o.\gamma_o$ is the removed rate for which infected individuals (symptomatic and non symptomatic) recover or die, leaving the infected groups $I_D(t)$ *and* $I_G(t)$. The accumulated SARS-Cov-2 confirmed cases are obtained from:

$$C(t) = \alpha_S.\int_0^t[\sigma_D.E_D(\tau) + \sigma_G.E_G(\tau)].d\tau \quad (7)$$

where $\alpha_S$ is the estimated fraction of symptomatic individuals. Major assumptions for this model are respectively the incubation rates: $\sigma_D$ and $\sigma_G$ for D and G SARS-CoV-2 exposed hosts.

A key parameter in deterministic transmission models is the basic reproductive number $R_o$, which is quantified by both, the pathogen and the particular population in which it circulates. Thus, a single pathogen, like the SARS-CoV-2, will have different $R_o$ values depending on the characteristics and transmission dynamics of the population experiencing the outbreak. When infection is spreading through a population that may be partially immune, it has been suggested to use an effective reproductive number $R(t)$. Serial interval (SI) is an essential metric for estimating both $R_o$, and $R(t)$, to predict disease trends, interventions and health care demands. SI depends on the pathogen incubation period which quantifies the biological process of relevant virus mutation, disease progression and tends to follow distributions resulting from genetic differences.

The methodology to estimate $R_o$ follows: The exponential growth rate of the epidemic, $r$ was obtained from the early stages of the epidemic, before NPIs were applied. The growth rate $r = 0.31 \pm 0.02$ of infected people was estimated applying the Levenberg-Marquardt method[10] to data of symptomatic infected people (Fig. 1), during the first 15 days of exponential growth according to the expression $I(t) = I_o.exp(-r.t)$.

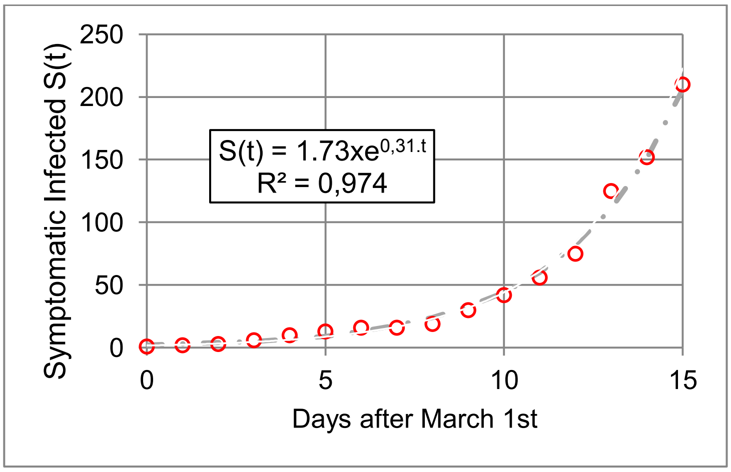


*Figure 1. Exponential fitting of the initial growth of symptomatic infect people. The growth rate r = 0.31 ± 0.02 was estimated applying the Levenberg-Marquardt method.*

The time period required to double the number of symptomatic cases is straightforward given by $\ln(2)/r = 2.3 \pm 0.1$ days. The basic Reproductive Number $R_o = 2.53 \pm 0.09$ was estimated according to; "In an epidemic, driven by human-to-human transmission, whereas growing exponentially, in a deterministic manner, the incidence I(t) can be described by the Renewal Equation", or the Lotka–Euler equation[11,12]:

$$I(t) = \int_0^\infty I(t-\tau)\beta(\tau)d\tau \quad (8)$$

where $\beta(\tau)$ is the mean rate at which an individual infects others a time after being infected itself. Substituting into Eqn. (8) an exponentially growing incidence, $I(t) = I_o.\exp(r.t)$, $I_o = 1$, gives the condition,

$$1 = \int_0^\infty e^{-\tau.r}\beta(\tau)d\tau \quad (9)$$

where $$\beta(\tau) = R_o.\omega(\tau) \quad (10)$$

ϖ(τ) is the generation time distribution, i.e. the probability density function for the time between an individual becoming infected and their subsequent onward transmission events. Ro is the basic reproduction number. If the exponential growth rate r and the generation time distribution ϖ(τ) have been estimated, Ro is readily determined from Eqn. (9), as

$$R_o^{-1} = \int_0^{\infty} e^{-r.\tau}.\,\omega_n(\tau,\lambda,\kappa).\,d\tau \qquad (11)$$

The term that appears in the right-hand side of this equation is the Laplace transform of the integrand function. More specifically, it is known as the Momentum Generating Function of this distribution. A normalized Gamma generation distribution is adopted (Eqns. 12, 13 and 14), with mean = k.θ = 3.6, SD = $k.\theta^2$ = 4.8, and peak value at 2.3 days (Fig. 2).

$$\omega_n(\tau,\lambda,\kappa) := \frac{\omega(\tau,\lambda,\kappa)}{I_W} \qquad (12)$$

$$\omega(\tau,\lambda,\kappa) := \left(\frac{\kappa}{\lambda}\right)\cdot\left(\frac{\tau}{\lambda}\right)^{(\kappa-1)}\cdot e^{-\left(\frac{\tau}{\lambda}\right)\cdot\kappa} \qquad (13)$$

$$I_W := \int_0^{200} \omega(\tau,\lambda,\kappa)\,d\tau \qquad (14)$$

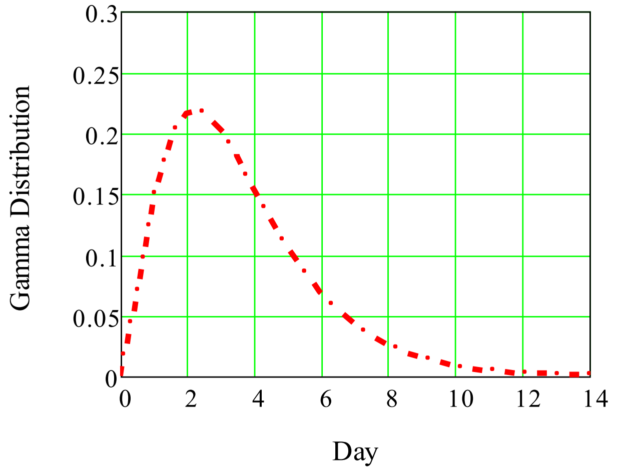


*Figure 2. Distribution of generation times or SI is presented. Data was described by the Gamma distribution with mean = 3.6 days, SD =4.8 days, and peak value at day 2.3. Dates of symptom onset with intervals of exposure for both source and recipient (when available) were collected[11] in order to select the best distribution.*

The real-time transmissibility of an infectious disease is better characterized by an instantaneous reproduction number *R(t)* defined as the expected number of secondary infections caused by an infector within a short time window. Equivalently *R(t)* can be expressed as the transmission rate *β(t)* divided by the rate $\gamma_o$ at which infected people recover or die. Mitigation policies aim to control the outbreak reducing the *R(t)* value. A number of methods are available to estimate effective reproduction numbers during epidemics. With this aim, a method for estimating *R(t)* using branching processes was developed. It relies on two inputs: a disease incidence time series (the numbers of new observed cases at successive times) and an estimate of the distribution of SI. In this study, during the outbreak in Brazil, *R(t)* was estimated following the procedure available on-line.[9] The result time-varying *R(t)* number obtained is presented in Fig. 3.

#### Supplementary Material

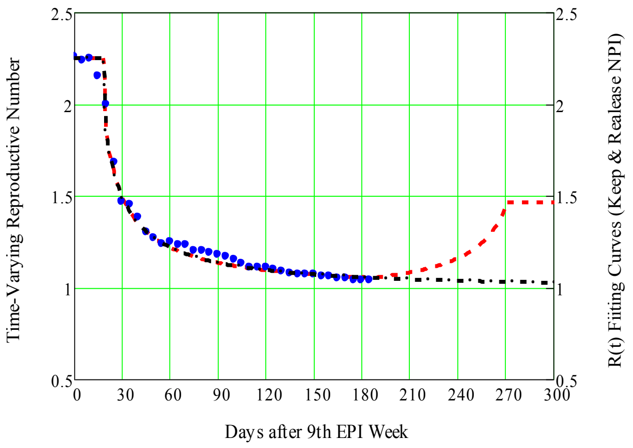


*Figure 3. Estimated R(t) values following the available on-line procedure[9] (solid blue circles). Two fitting functions are selected to model the evolution of R(t) in different scenarios. This first fitting is shown (dash-and-dot brown trace) as continuous approach of R(t) to its unit value. On a different scenario, aiming to model the second COVID-19 wave spreading in Brazil, a second fitting is proposed (dash red trace).*

Assuming to the end of 2020 the preservation of all the social NPI, as social distancing, mask wearing, gloves, hand wash, among others, a first fitting function is selected to model a continuous approach of R(t) to its unit value. This first fitting is shown in Fig.3 as a dash-and-dot black trace. On a different scenario, aiming to model the second COVID-19 wave spreading in Brazil during the second half of 2020, a second fitting is proposed. This second fitting is included in Fig. 3 as a dash red trace. In this scenario, a progressive release of all NPI is adopted leading *R(t)* to return to 65% of its initial value $R_o$. Both scenarios will be discussed latter.

The set of ODE (15) was numerically solved using PTC MathCad, and assuming the following initial conditions: $S(0) = N - E_o$, $\beta(0) = 1.265$, $E_D(0) = 0.38E_o$, $E_G(0) = 0.62E_o$, $I_D(0) = I_G(0) = R(0) = 0$. It is assumed that both D and G groups share the same *R(t)* function. In short, $N$, $\sigma_D = 1/4$, $\sigma_G = 2\sigma_D$ , and $\beta(0)$ are the only fitting parameters to data. The susceptible number $N=60$ Million (28.7% of the Brazilian population) is chosen as the minimum number of susceptible hosts to account for the accumulated confirmed cases in the study period.

$$\frac{d}{dt}S(t) = -\beta(t)\cdot(I_D(t) + I_G(t))\cdot\frac{S(t)}{N} \qquad \frac{d}{dt}I_D(t) = -\gamma_o\cdot I_D(t) + \sigma_D\cdot E_D(t)$$

$$\frac{d}{dt}E_D(t) = \beta(t)\cdot(I_D(t))\cdot\frac{S(t)}{N} - (\sigma_D\cdot E_D(t)) \qquad \frac{d}{dt}I_G(t) = -\gamma_o\cdot I_G(t) + \sigma_G\cdot E_G(t)$$

$$\frac{d}{dt}E_G(t) = \beta(t)\cdot(I_G(t))\cdot\frac{S(t)}{N} - (\sigma_G\cdot E_G(t)) \qquad \frac{d}{dt}R(t) = \gamma_o\cdot(I_D(t) + I_G(t))$$

(15)

$$\begin{pmatrix} SS \\ EE_D \\ EE_G \\ II_D \\ II_G \\ RR \end{pmatrix} := \text{Odesolve}\left[\begin{pmatrix} S \\ E_D \\ E_G \\ I_D \\ I_G \\ R \end{pmatrix}, t, t_F\right]$$

The total number of cases or infected hosts (symptomatic and non-symptomatic) is given by Eqn. 16. The numbers of reported symptomatic individuals, and fatalities are estimated from Eqn. 17:

### Supplementary Material

$$\text{cases}(u) := \int_0^u \left(\sigma_D \cdot EE_D(t) + \sigma_G \cdot EE_G(t)\right) dt \tag{16}$$

$$C(u) := \alpha_S \cdot \text{cases}(u-4) \qquad \text{Fatal}(u) := c(u) \cdot C(u) \tag{17}$$

Where c(u) is determined from daily reported official data on cases and fatalities, and analytically extended (Fig. 4) to forecast future deaths. Fatalities were modeled as a function of the confirmed cases C(u), and this dependence, obtained from official reported cases and deaths, is not linear neither monotonic as discussed in the main article.

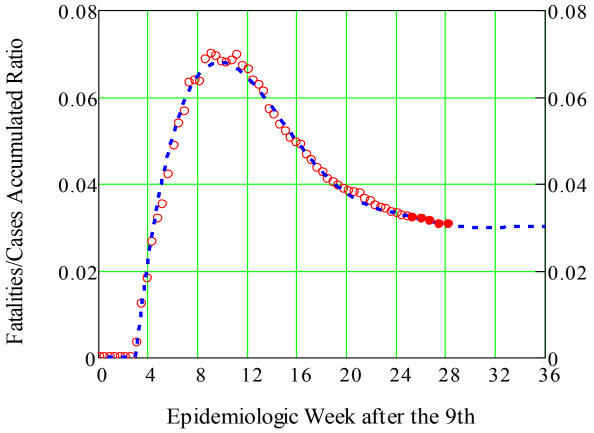


*Figure 4. Time dependence of the ratio Fatalities/(Confirmed Cases). This ratio presents a time-dependence that is not linear neither monotonic. The non-zero ratio of deaths/(confirmed cases) by COVID-19 (open red circles), begins three weeks after the first reported case, rises monotonically, after roughly seven weeks reaches a maximum of 7%, and drops to a ratio around 2.6 %.*

Official Brazilian data[13] from February 29 to August 19 2020 (EPI week # 9), was considered in order to estimate the main parameters that govern the dynamics established by Eqns. (1) to (7) Model parameters were estimated by minimizing the mean squared quadratic errors, where M = 176 days. The Standard Deviation (SD - log scale) between data and model (Eqn. 18) is only 7% over six orders of magnitude, as presented by Fig. 5.

$$\text{RMS} := \left[\left(\frac{1}{M}\right) \cdot \sum_{i=6}^{M} \left(\log(C(i)) - \log(\text{Cases}_{0,i})\right)^2\right]^{\frac{1}{2}} = 0.09 \qquad \text{SD} := \left(\frac{1}{M}\right) \cdot \sum_{i=6}^{M} \left|\log(C(i)) - \log(\text{Cases}_{0,i})\right| = 0.07 \tag{18}$$

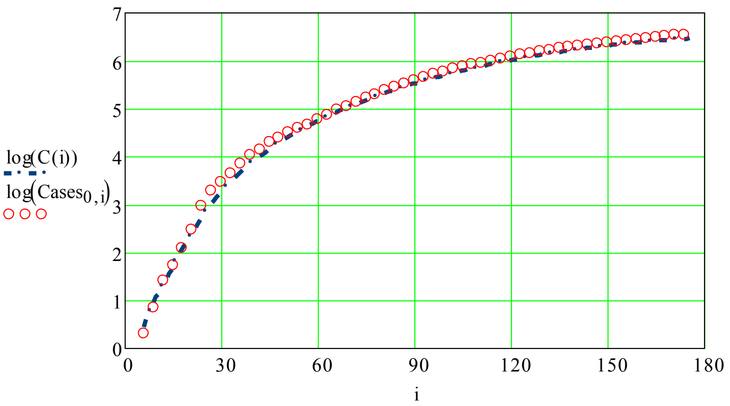


*Figure 5. The Standard Deviation (SD - log scale) between data and model (Eqn. 18) is only 7% over six orders of magnitude.*

#### Supplementary Material

According to the results obtained solving the set of ODE (15) and shown by Fig. 6, the fraction of depleted susceptible SS(t)/SS(0), preserving the NPIs, saturates bellow 15 %. On the other hand, the progressive release of the NPIs allowing R(t) to return and reach 65% of $R_o$ by the end of 2020, depletes the susceptible individuals in roughly 35 %.

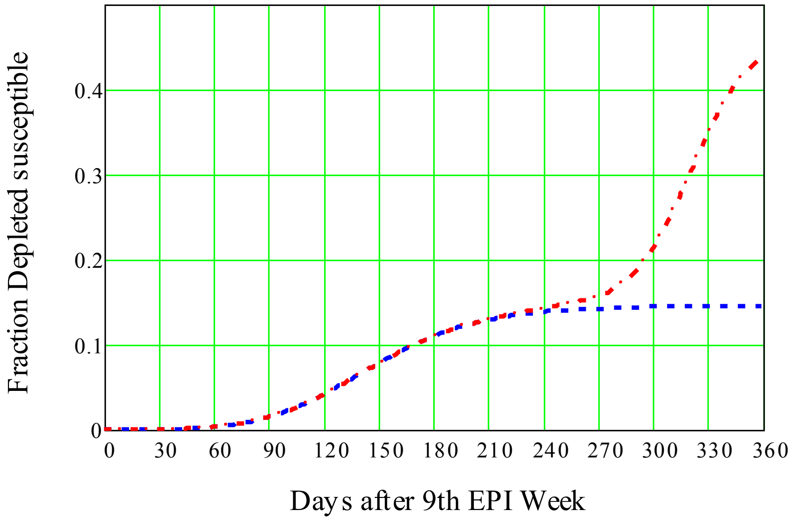


*Figure 6. The fraction of depleted susceptible SS(t)/SS(0), preserving the NPIs, saturates bellow 15 % (dashed blue trace). On the other hand, a progressive release of the NPIs, allowing R(t) to return and reach 65% of $R_o$ by the end of 2020, depletes the susceptible individuals in roughly 35 %.*

Regarding the original SARS-CoV-2 D-form and its G-variant, the only distinction in modeling is their own incubation rates. It is assumed that both forms produce the same disease fatalities, and share the same instantaneous reproductive number. The infected D and G hosts are given by Eqn. 19:

$$I_g(u) := \int_0^u \gamma_o \cdot II_G(t)\, dt \qquad I_d(u) := \int_0^u \gamma_o \cdot II_D(t)\, dt \qquad (19)$$

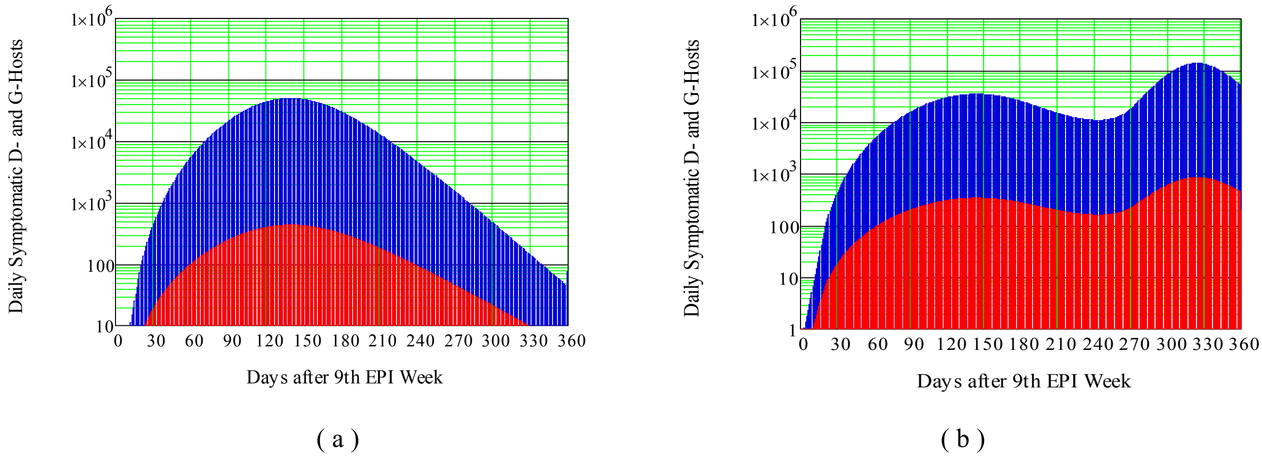


*Figure 7. Estimated evolutions of symptomatic infected host to SARS-CoV-2 original D-form and G-variant. Red and blue shades present respectively the evolution of D and G exposed hosts to SARS-CoV-2. (a) Preserving NPIs as indicated in Fig.3. (b) Progressive release of NPIs as reported world wise during the last quarter of 2020, as modeled in Fig.3.*

Figure 7 presents daily evolution of symptomatic infected host to the original D-form and its G-variant according to model estimative. These results confirm the prevalence of the G-variant form in COVID-19 pandemic as globally predicted, and reported for South America. They also point out to the presence of a second outbreak by the end of 2020 as a result of the release of NPIs across the country. These previsions suggest reinforcing the NPIs, and continuing the surveillance of SARS-CoV-2 mutations to support development of immunological interventions.

#### Conclusions

The results presented here demonstrate that this method can be employed to describe pandemic dynamics beyond the Brazilian cohort study case. The epidemic in Brazil was selected as a first study case. Such results can be functional to a more quantitative understanding of future pandemics, and this methodology can be applied worldwide to predict forecasts of the outbreak in any infected country. An important conclusion worth to point out follows. Despite the social and mental commotion that COVID-19 has imposed so far, most of the Brazilian and the world populations are still susceptible to SARS-CoV-2 infection. By the end of 2020, a fraction in the range of 15–35 percentages of susceptible Brazilian individuals is predicted to be depleted. Sufficient depletion of susceptibility (by NPIs or not) has to be achieved to weaken the global dynamics spread.

#### Acknowledgments

The author acknowledges the financial support from FUNTTEL. Financial grant # 01.16.0053.01 FINEP/MCTI received from the Brazilian Ministry of Science, Communications, and Innovation. This paper reflects only the author's views and the Agencies are not responsible for any use that may be made of the information contained therein. Thanks to Dr Kiyoshi Yoneda and Dr Roberto Panepucci for enlighten suggestions.
